# Clonal spread of *Plasmodium falciparum* candidate artemisinin partial resistance *Kelch13* 622I mutation and co-occurrence with *pfhrp2/3* deletions in Ethiopia

**DOI:** 10.1101/2023.03.02.23286711

**Authors:** Abebe A. Fola, Sindew M. Feleke, Hussein Mohammed, Bokretsion G. Brhane, Christopher M. Hennelly, Ashenafi Assefa, Rebecca M. Crudal, Emily Reichert, Jonathan J. Juliano, Jane Cunningham, Hassen Mamo, Hiwot Solomon, Geremew Tasew, Beyene Petros, Jonathan B Parr, Jeffrey A. Bailey

## Abstract

The emergence and spread of drug- and diagnostic-resistant *Plasmodium falciparum* are major impediments to malaria control and elimination. We deep sequenced known drug resistance mutations and other informative loci across the genome of 609 samples collected during a study across three regions of Ethiopia. We found that 8.0% (95% CI 7.0-9.0) of malaria cases were caused by *P. falciparum* carrying the candidate artemisinin partial-resistance *K13* 622I mutation, which occurred less commonly in diagnostic-resistant *pfhrp2/3-*deleted than normal non-deleted parasites (*p*=0.03). Identity-by-descent analysis showed that 622I parasites were significantly more related than wild-type (p<0.001), consistent with recent expansion and spread. *Pfhrp2/3-*deleted parasites were also highly related, with evidence of clonal transmissions at the district level. Parasites carrying both *pfhrp2/3* deletion and 622I mutation were observed in some sites. These findings raise concern for future spread of combined drug- and diagnostic-resistant parasites and warrant close monitoring.

## Introduction

Despite intensified malaria control efforts, progress toward elimination has stalled in recent years. Malaria cases increased across many endemic countries in Africa, where 96% of malaria deaths occur primarily due to *Plasmodium falciparum*^*1*^. The World Health Organization (WHO) recommends artemisinin-combination therapies (ACTs), such as artemether-lumefantrine (AL) or artesunate-amodiaquine (AS-AQ), as the first-line treatments for uncomplicated *P. falciparum* malaria^2^. However, the parasite has evolved drug resistance to most available antimalarial drugs^3,4^ and history demonstrates that resistant strains can rapidly spread^5,6^. Since 2008, *P. falciparum* parasites resistant to first-line ACTs have emerged in Southeast Asia^7,8^ and have spread to neighboring regions^9,10^.

Increasing reports across Africa indicate reduced efficacy of artemisinins, with slowed clearance times and increased recrudescences^11–14^. Mutations in *kelch13* (*K13*) associated with partial resistance to artemisinins have now also been reported in Uganda, Tanzania and Rwanda^15–17^. In addition, parasites undetectable by widely used *P. falciparum* rapid diagnostic tests (RDTs), owing to deletion mutations of the histidine-rich proteins 2 and 3 (*pfhrp2/3*) genes, have emerged in the Horn of Africa^18–20^. Together, these mutations threaten both components of existing test-and-treat programs, as co-occurrence of *pfhrp2/3* deletions and *K13* mutations would yield parasites resistant to both diagnosis and treatment. Improved understanding of how these mutations emerge, interact, and spread is critical to the success of future malaria control and elimination efforts across Africa.

In Ethiopia, malaria is endemic across 75% of the country, with 65% of the population at risk^21^. Over five million episodes of malaria occur each year, but transmission is highly heterogeneous and seasonal^22^. Prompt diagnosis and treatment with efficacious drugs is a cornerstone of the malaria program^23^. ACTs are first-line treatment for uncomplicated falciparum malaria since 2004 throughout the country. However, chloroquine (CQ) is still widely used and efficacious for cases of endemic vivax malaria^24^. Artemether-lumefantrine remains highly efficacious^25^, though detection of the candidate artemisinin resistance *K13* 622I mutation in northern Ethiopia^26,27^ and high prevalence of residual submicroscopic parasitemia following ACT treatment in recent studies raises concern^12,25^.

Countries such as Ethiopia provide important opportunities to investigate the impact of drug and diagnostic resistance mutations and how they may co-evolve and spread. To our knowledge, there are no published studies addressing the prevalence of drug resistance mutations among *pfhrp2/3-*deleted versus non-deleted strains, nor their transmission patterns. We sought to bridge this knowledge gap by conducting a comparative genomic analysis of drug resistance among *pfhrp2/3-*deleted and non-deleted parasites collected across three regions of Ethiopia. Using molecular inversion probe (MIP) sequencing for highly-multiplexed targeted genotyping^28,29^, we determine the prevalence of key drug resistance mutations and detect spatial patterns of related 622I mutant and *pfhrp2/3*-deleted *P. falciparum* strains. We demonstrate a high prevalence of the 622I mutation across three regions and co-occurrence with *pfhrp2/3* deletion in Ethiopia as well as clonal transmission of *pfhrp2/3* deleted parasites at district level.

## Results

### MIP sequence data filtering and complexity of infections estimation

A total of 920 samples previously genotyped and MIP sequenced for *pfhrp2/3* deletions from three regions of Ethiopia (Amhara = 598, Gambella = 83, Tigray = 239) (**Supplementary Figure S1)** were included in this analysis, representing dried blood spots taken from a subset of the overall series of 2637 malaria cases (Amhara = 1336, Gambella = 622, Tigray = 679) (**Table S1**). Samples had been collected from rural areas in 12 districts as part of a large *pfhrp2/3* deletion survey of those 12,572 study participants (56% male, 44% female, age ranges 0 and 99 years) presenting with clinical signs and symptoms of malaria^18^. For this study, all samples were further MIP captured and sequenced using both i) a drug resistance panel comprising 814 probes designed to target mutations and genes associated with antimalarial resistance and ii) a genome-wide SNP panel comprising 1832 probes designed for assessment of parasite relatedness and connectivity (**Supplementary Data 1 and 2**). Parasite densities across samples ranged from 3 to 138,447 parasites/µl with median parasitaemia of 1,411 parasites/µl (**Supplementary Figure S2A**); as expected, MIP sequencing coverage was parasite density-dependent (**Supplementary Figure S2B**). All resistance genotypes with sufficient depth and quality were included in downstream analysis. After filtering for sample missingness and removing loci with low coverage (**Supplementary Figure S3)**, 609 samples and 1395 SNPs from the genome-wide panel (**Supplementary Figure S4, Supplementary Data 3 and 4**) were included in downstream relatedness analysis.

Using filtered genome-wide SNPs, we calculated complexity of infection (COI) and adjusted for the relative proportion of DBS sampled from participants with discordant vs. concordant RDT results since the parent *pfhrp2/3* survey purposefully oversampled the former. We estimate that the majority (82.4%) of cases are monogenomic infections (COI = 1) (**Supplementary Figure S5, Table S1**), reflecting relatively low ongoing transmission in the study areas. Overall, COI per sample ranged from 1 to 4 with variability at the district level (**Supplementary Figure S5C**), consistent with heterogeneous malaria transmission at local scale.

### *K13* 622I mutation is prevalent in Ethiopia across all regions sampled

Analysis of the drug-resistance markers revealed a high prevalence (8.0%, [95% confidence interval (CI) 7.0-9.0]) of samples expected to carry the WHO candidate artemisinin partial resistance mutation 622I within the propeller domain of *K13*. The 622I mutation had only been previously described in Africa at a single site in Amhara near the Sudan border in 2014 at 2.4% prevalence^26^. Our results confirmed parasites with 622I in all 3 regions surveyed as well as all 12 districts (**Figure 1A)**. Highest prevalence was observed in Amhara (9.8%, [95% CI 8.2-11.4]) in the northwest near the Sudan border, followed by Tigray (8.4%, [95% CI 6.2-10.5]) near the Eritrea border, and Gambella (3.6%, [95% CI 2.1-4.8]) bordering South Sudan, however, there was high spatial heterogeneity at the district level and within regions (**Table S1**). An additional 8 non-synonymous mutations were identified across the *K13* gene at low frequencies (<3%) except for K189T (44.4%), which is frequently observed in Africa and not associated with resistance (**Figure 1B**). None of the other mutations were WHO-validated or candidate artemisinin partial resistance mutations, and only two (*K13* E401Q and E433D) fell within the propeller region (**Figure 1B, bottom panel, Supplementary Table S1**). To gain insight into relative fitness of 622I, we compared within-sample allele proportions in mixed mutant and wild-type infections (n =16). On average, wild-type parasites occured at relatively higher proportions (mean = 0.59) compared with 622I mutant parasites (mean = 0.41) (Mann-Whitney *p* = 0.025) in participants infected by more than one strain, suggesting lower fitness of mutant strains. The power of this analysis was limited as polygenomic infections were rare in this study but is consistent with competitive blood stage fitness costs.

**Figure 1.**
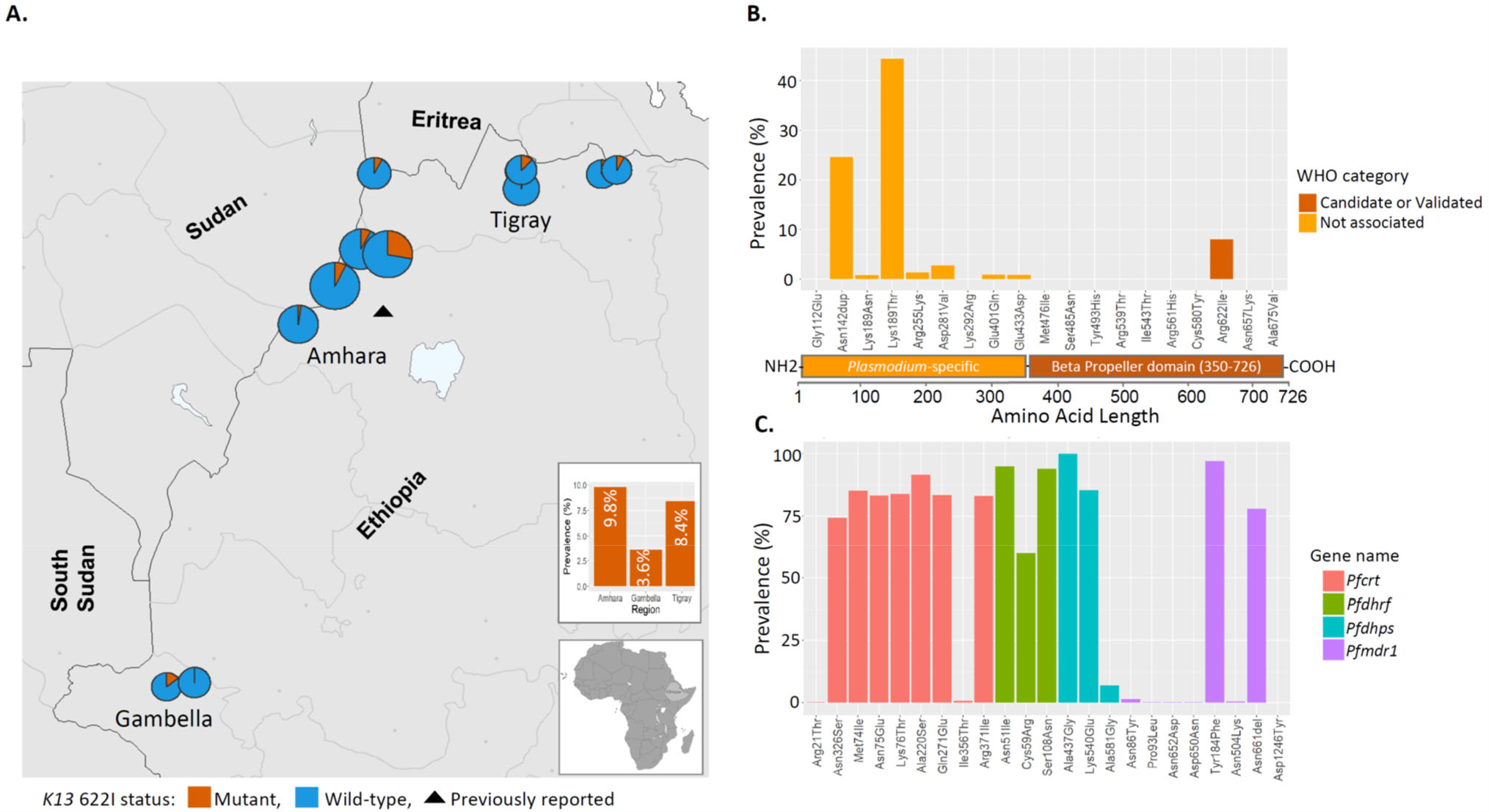
Prevalence of *K13* and key drug-resistance mutations in Ethiopia. **A)** Spatial distribution of *K13* 622I mutation at the district (pie charts) and regional (bar plot) levels. Colors indicate mutation status, and pie chart size is proportional to sample size per district. The black triangle indicates the location where *K13* 622I mutation was reported previously. **B)** Prevalence of nonsynonymous mutations across the *K13* gene, colored according to WHO ACT resistance marker category. *K13* gene annotation shows 1-350 amino-acid residues in the poorly conserved *Plasmodium-*specific region, and 350-726 residues in the beta propeller domain where validated resistance mutations are located. **C)** Prevalence of mutations across four key *P. falciparum* genes associated with commonly used antimalarial drugs (colors).

### *K13* 622I mutant parasites carry background mutations that may augment ACT resistance

In addition to *K13* mutations, we found a number of key mutations in other *P. falciparum* genes associated with resistance to different antimalarial drugs (**Figure 1C, Table S2**), including ACT partner drugs. Mutations in the *P. falciparum* multidrug resistance gene 1 (*pfmdr1*), particularly isolates that carry the NFD haplotype (N86Y (wild), Y184F (mutant), and D1246Y (wild)) have been associated with decreased sensitivity to lumefantrine^30^. Overall, 83% of samples carry the NFD haplotype (**Figure 2)**, and 98% (60/61) of 622I mutant parasites carry *pfmdr1* NFD haplotypes. Although this difference was not significant (Fisher’s exact *p* = 0.34), the presence of 622I mutant parasites with *pfmdr1* NFD haplotypes raises questions about how the genetic background of 622I influences ACT efficacy in Ethiopia. We also investigated other mutations previously identified as backbone loci on which artemisinin partial resistance associated *K13* mutations are most likely to arise or could augment ACT resistance^31^. No parasites sampled in this study carried such background mutations (*pffd*-D193Y, *pfcrt*-I356T, *pfarps*-V127M and *pfmdr2*-T484I), with the exception of *pfcrt*-N326S, which 98% of *K13* 622I and 81% of wild-type parasites carried (Fisher’s exact *p* < 0.001) (**Figure 2**). The co-occurrences of 622I with the *pfmdr1* NFD haplotype and *pfcrt-*N326S raise concern about the efficacy of both artemisinin and partner drugs like lumefantrine in Ethiopia. We also observed drug-resistance mutations in other genes (**Table S3**), with high prevalence and some spatial heterogeneity in the distribution of mutations associated with sulfadoxine-pyrimethamine (SP) resistance **(Supplementary Figure S6**).

**Figure 2.**
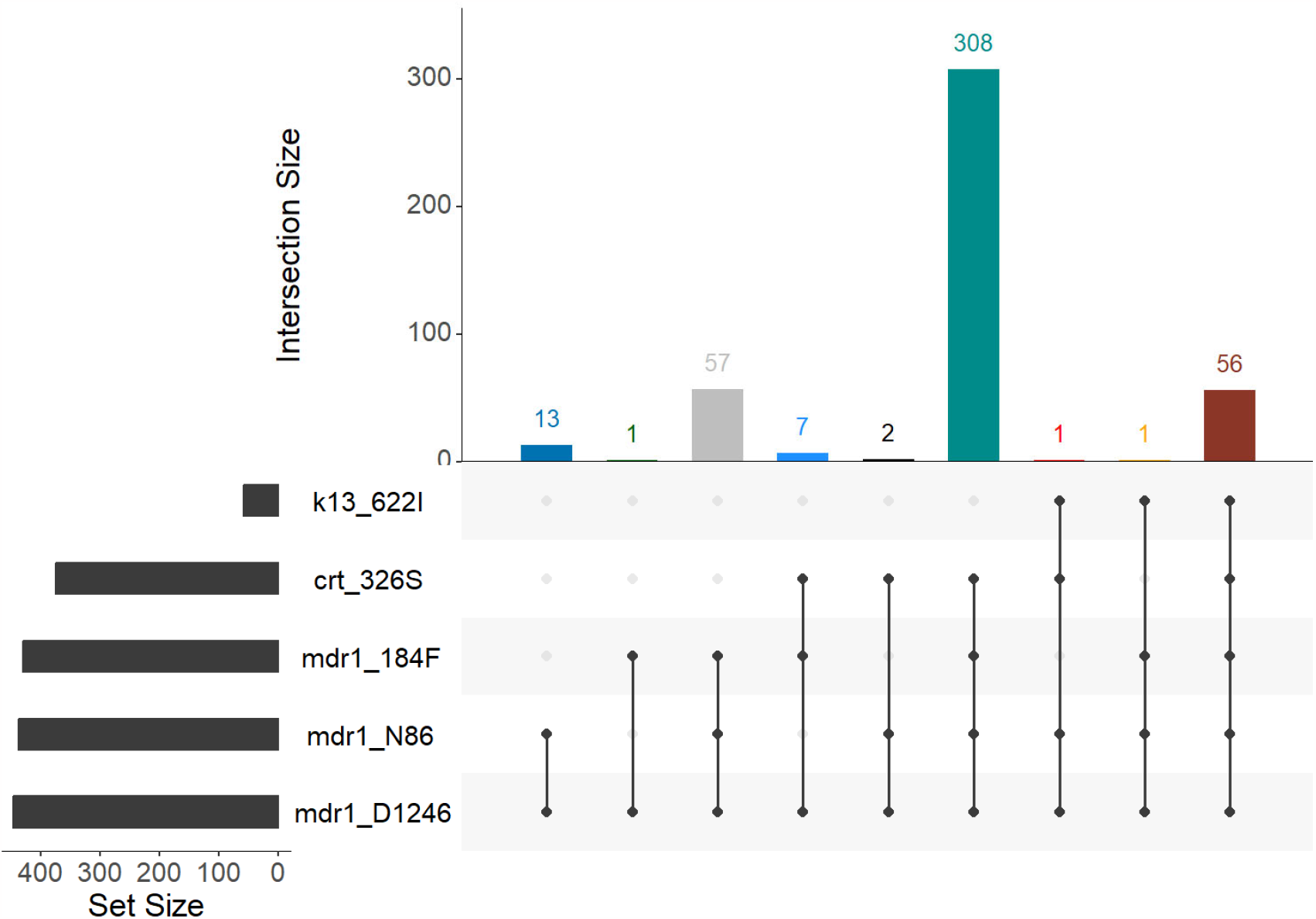
Frequency of key drug resistance mutations. The number of times each combination of mutations was observed is displayed, including *K13* 622I *pfmdr1* N86 (wild), 184F (mutant), and D1246 (wild)); and *pfcrt* genes. Results from monogenomic or the dominant haplotype in polygenomic infections are shown. Only samples with complete genotypes across all loci are shown.

### Co-occurrence of drug-resistance mutations and *pfhrp2/3* deletions

Overall, the *K13* 622I mutation is more common among *pfhrp2/3* non-deleted parasites (26/223, 11.6%) than *pfhrp2/3* double*-*deleted parasites (5/110, 4.5%), though not significantly (Fisher’s exact *p* = 0.07). However, higher mean prevalence of 622I mutation is observed among *pfhrp2/3* non-deleted parasites at the district level (T-test *p* = 0.03) (**Figure 3A**), which could be consistent with deleterious effects from the combination and/or independent origins with slow intermixing. We repeated this analysis using permutation by randomly reassigning double- and non-deleted groups and took the mean difference of these new groups. The permutation analysis shows -8.7% mean difference (F-statistic *p* = 0.02) in prevalence of 622I among *pfhrp2/3-*deleted vs. non-deleted parasites, suggesting patients infected by double-deleted parasites are more likely misdiagnosed and less likely receive ACTs according to the country test and treat policy that result in less ACT drug pressure. We observed a negative correlation between these mutations at the level of the individual collection sites, suggesting different sites generally harbor one mutation or the other at high frequency. However, we observed a small number (n = 5) of parasites with both 622I mutation and *pfhrp2/3* deletion in sites where mutation or deletion frequency is high (**Figure 3B**), confirming that recombination between parasites with these mutations is possible. Interestingly, 622I is more common among *pfhrp3-*deleted parasites (29/169, 17.2%) compared to wildtype *pfhrp2/3* non-deleted (26/223, 11.2%) but the difference was not statistically significant (Chi-square *p* = 0.23).

**Figure 3.**
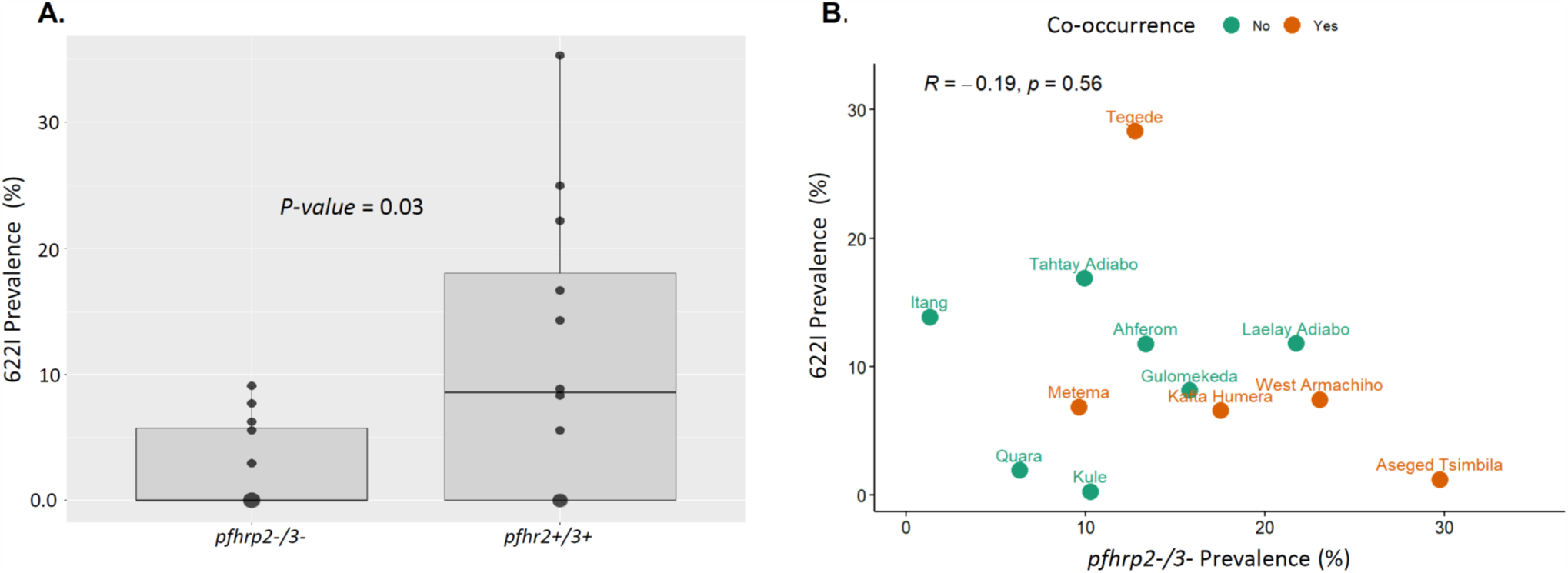
*K13* 622I mutation among *pfhrp2/3-*deleted and *pfhrp2/3 non-*deleted parasite populations. **A**) Comparison of mean *K13* 622I mutation prevalence between *pfhrp2/3* double and *pfhrp2/3* non-deleted parasite populations by district across three regions in Ethiopia. **B**) Relationship between *pfhrp2/3* double*-* deleted parasite prevalence and *K13* 622I mutation prevalence by district. Prevalence estimates are weighted. Orange points represent districts where a parasite harboring both *pfhrp2-/3-* deletion and *K13* 622I mutations are observed.

We also examined co-occurrence of *pfhrp2/3* deletions and other drug resistance mutations, particularly *pfcrt* mutations as most *pfhrp2/3* deletion reports to-date have emerged in areas where *P. vivax* and *P. falciparum* are sympatric and chloroquine is used to treat vivax malaria^32^. We observed overall high prevalence (median 84% across districts) of *pfcrt* mutations (codon 74-76) (**Supplementary Figure S7A**). The prevalence of *pfcrt-*K76T mutation was greater among *pfhrp2/3*-deleted (96.3%) compared to non-deleted (73.8%) parasites, but the difference was not statistically significant (Chi-square *p* = 0.15, **Supplementary Figure S7B**). This finding suggests patients infected by *pfhrp2/3*-deleted parasites may be more often exposed to chloroquine.

### Population structure of *K13* 622I and *pfhrp2/3-*deleted *P. falciparum* in Ethiopia

We investigated genetic population structure using principal component analysis (PCA), which revealed clustering of parasites by *K13* 622I mutation (PC1) and by *pfhrp2/3* deletion (PC2) status, but not by geography (**Figure 4, Supplementary Figure S8A**). Overall, 13.4% of variation in our dataset was explained by these first two principal components (**Supplementary Figure S8B**). Analysis of loading values did not reveal SNPs or genomic regions with disproportionate influence on the observed population structure (**Supplementary Figure S9)**. Genetic differentiation between populations is low overall (*F*_*st*_ range = 0.002 - 0.008), and lowest between Amhara and Tigray regions (*F*_*st*_ = 0.002) and highest between Gambella and Tigray regions (*F*_*st*_ = 0.008), followed by between Amhara and Gambella (*F*_*st*_ = 0.003).

**Figure 4.**
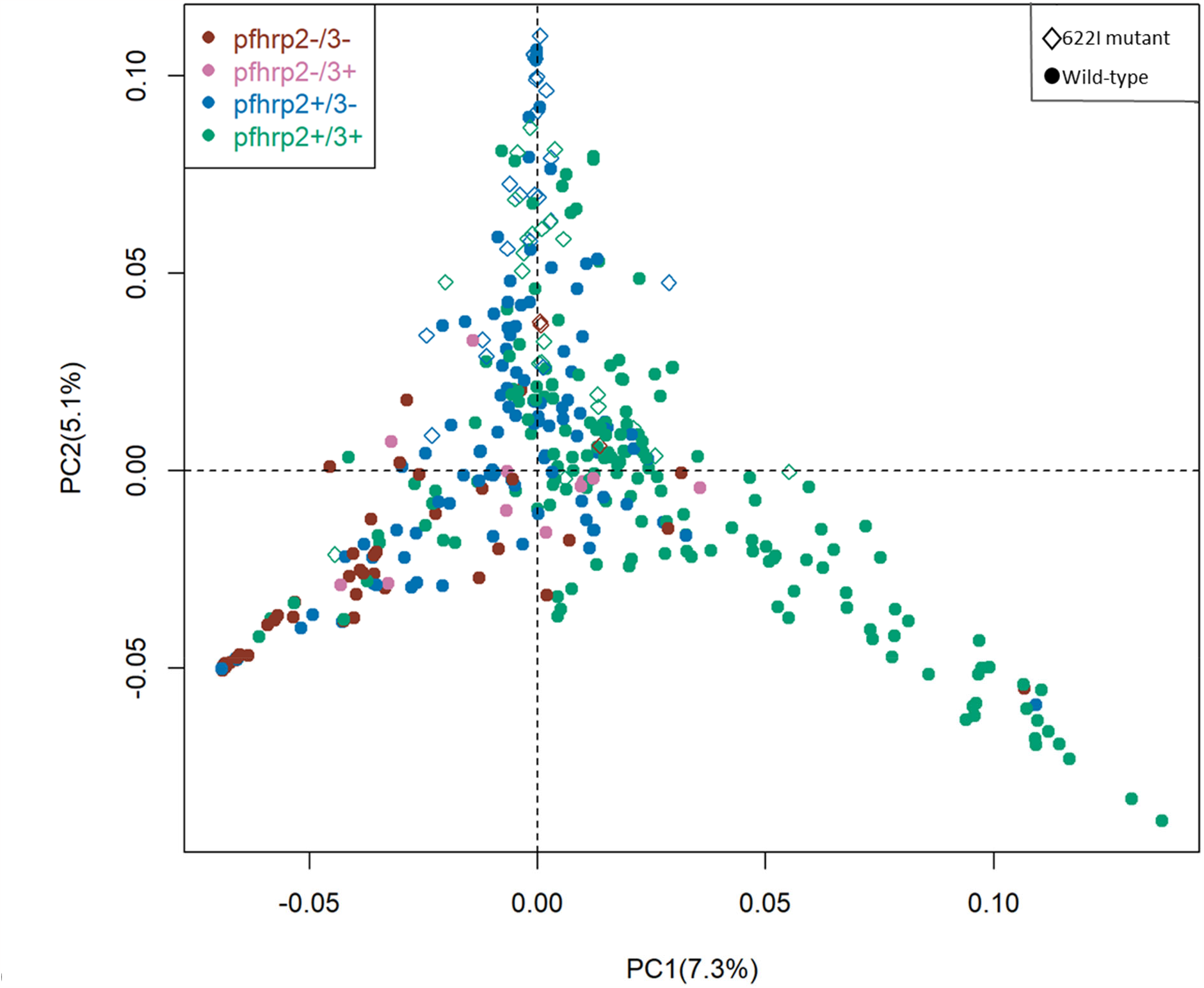
Principal component analysis of *P. falciparum* populations annotated by *K13* 622I and *pfhrp2/3* deletion genotypes. Colors indicate *pfhrp2/3* deletion status and shape indicates *K13* 622I mutation status. Percentage of variance explained by each principal component is presented.

### Genetic relatedness of *K13* 622I mutant and *pfhrp2/3-*deleted parasites

IBD analysis revealed evidence of recent clonal transmission and spread of *K13* 622I parasites. Overall, 10.6% of pairs are highly-related (IBD ≥ 0.25, half siblings) (**Figure 5A**). We observe a tailed distribution of highly related parasite pairs, with 26.6% of pairwise comparisons sharing their genome with an IBD value of ≥ 0.05. Comparing *K13* 622I mutant and wild-type parasites, we find significantly higher mean pairwise IBD sharing within *K13* 622I mutant populations (0.43 vs 0.08, respectively, Mann-Whitney *p* < 0.001) (**Figure 5B)**. Network analysis of highly related parasites (pairwise IBD ≥ 0.95) shows that 622I mutant parasites tend to form related clusters and pairs separate from wild-type parasites (**Figure 5C**), consistent with clonal transmissions of 622I parasite populations in Ethiopia. The majority of clonal parasites carrying 622I mutation originated from one district (Tegede) (**Figure 5D)**, likely illustrating an outbreak with rapid spread (**Table S1**).

**Figure 5.**
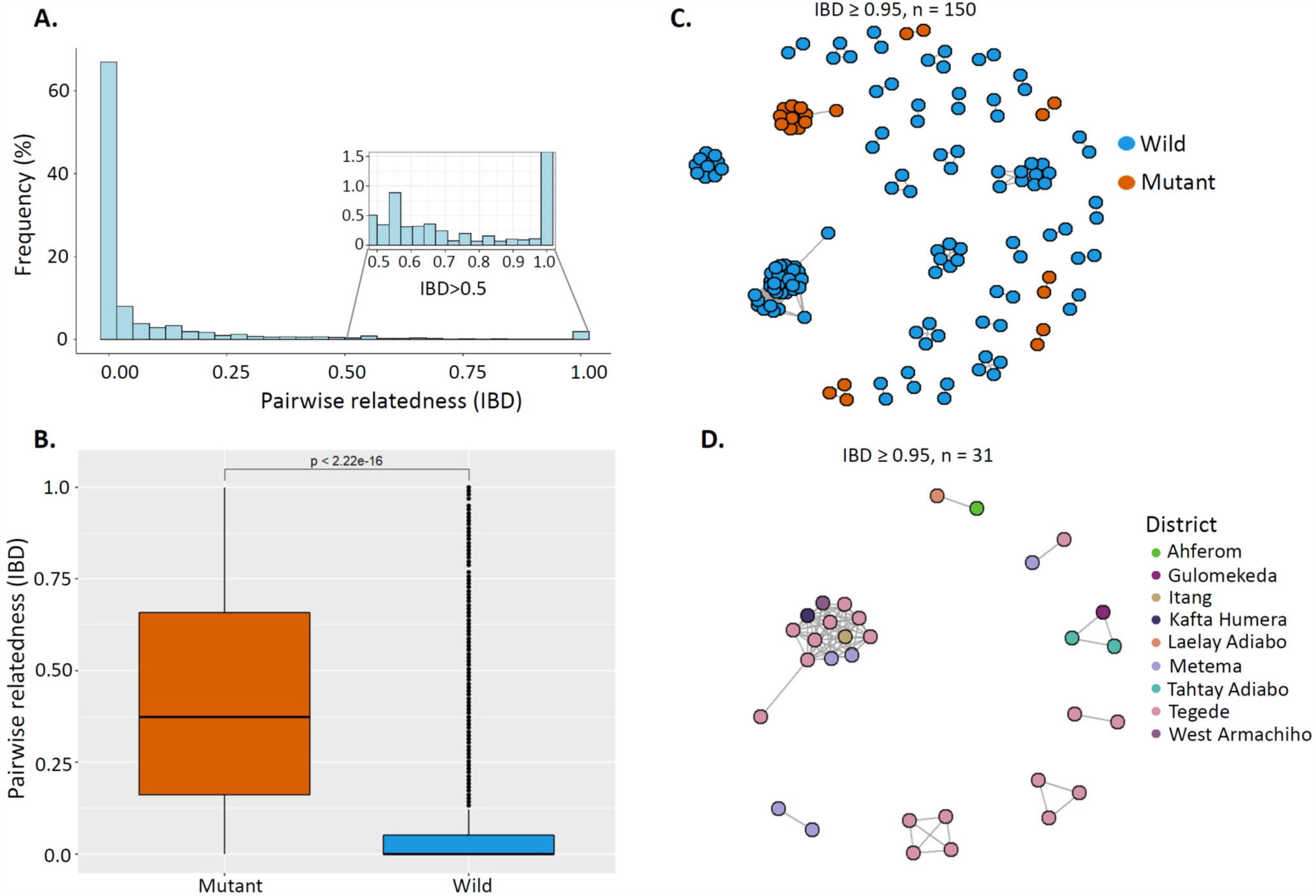
Pairwise IBD sharing and relatedness networks suggest clonal transmission and expansion of *K13* 622I parasites. **A)** Pairwise IBD sharing across all three regions of Ethiopia. The plot shows the probability that any two isolates are identical by descent, where the x-axis indicates IBD values ranging from 0-1 and y-axis indicates the frequency (%) isolates sharing IBD. The inset highlights highly related parasite pairs, with a heavy tail in the distribution and some highly related pairs of samples having IBD ≥ 0.95. **B)** Pairwise IBD sharing within parasites carrying *K13* 622I vs. wild-type. Boxes indicate the interquartile range, the line indicates the median, the whiskers show the 95% confidence intervals, and black dots show outlier values. *P value* determined using Mann-Whitney test is shown. **C)** Relatedness network of highly related parasite pairs sharing IBD ≥ 0.95. Colors correspond to *K13* 622I mutant and wild parasites. **D)** Relatedness network of only *K13* 622I parasite pairs sharing IBD ≥ 0.95 at the district level/local scale. Colors correspond to districts across three regions in Ethiopia. In both panels C and D, each node identifies a unique isolate, and an edge is drawn between two isolates if they share their genome above IBD ≥ 0.95. Isolates that do not share IBD ≥ 0.95 their genome with any other isolates are not shown.

*Pfhrp2/3-*deleted parasites also have higher relatedness than wild-type parasites, with significantly different pairwise IBD sharing (Kruskal-Wallis test *p* < 0.001) when comparing *pfhrp2/3* double-, single-, and non-deleted parasites (**Figure 6A**). Pairwise IBD sharing is highest among *pfhrp2/3* double-deleted parasites, with 43.7% of comparisons having IBD ≥ 0.25 (half siblings), compared to only 4.3% of *pfhrp2/3* non-deleted parasites. Network analysis of highly related isolates (IBD ≥ 0.95) revealed clustering by deletion status (**Figure 6B**) with district-level clustering of *pfhrp2/3-*double deleted parasites evident in Kule, Atse-Tshimbila and West-Armachiho (**Figure 6C**), a finding consistent with clonal spread of *pfhrp2/3-*double deleted parasites at the local scale.

**Figure 6.**
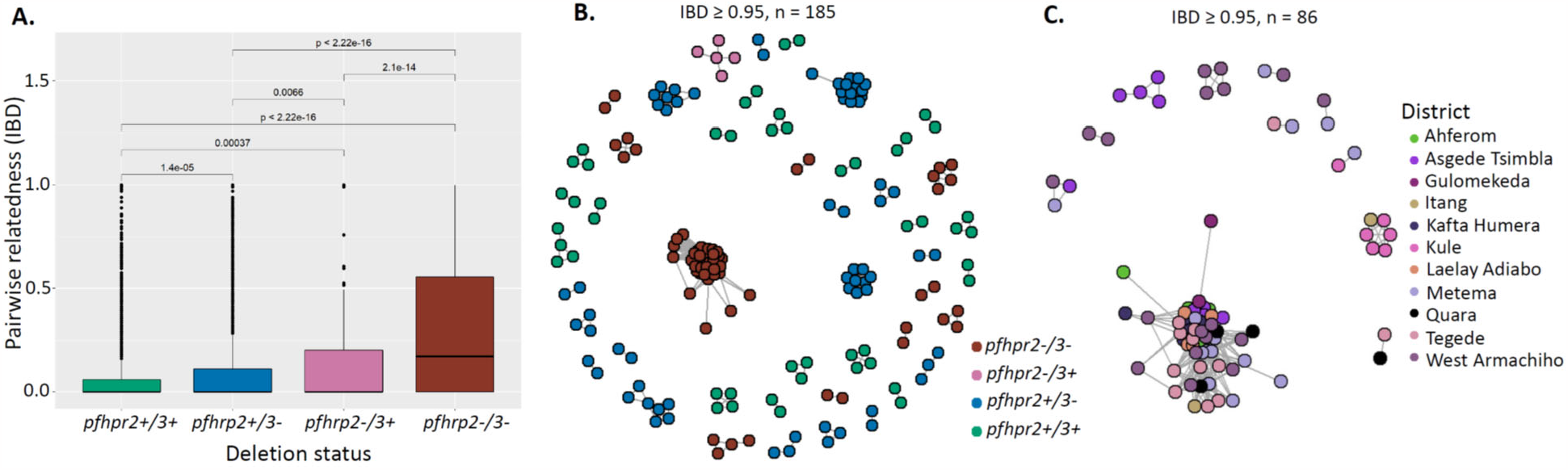
Pairwise IBD sharing and relatedness networks suggest independent emergence and clonal spread of *pfhrp2/3-*deleted parasites. **A)** Pairwise IBD sharing by *pfhrp2/3* deletion status. Boxes indicate the interquartile range, the line indicates the median, the whiskers show the 95% confidence intervals, and black dots show outlier values. *P* values were determined using the Kruskal-Wallis test. **B)** Relatedness network of highly related parasite pairs sharing IBD ≥ 0.95. Each node identifies a unique isolate, and an edge is drawn between two isolates if they share their genome with IBD ≥ 0.95. Isolates that do not share their genome IBD ≥ 0.95 with any other isolates are not shown. Color codes correspond to *pfhrp2/3* deletion status. **C)** Relatedness network of *pfhrp2/3* double-deleted parasite pairs with IBD ≥ 0.95 at district level/local scale. Colors correspond to districts across three regions of Ethiopia.

## Discussion

Our genetic analysis confirms a widely prevalent WHO candidate artemisinin partial resistance *K13* 622I mutation across three regions of Ethiopia and suggests recent clonal spread. We observe low levels of polyclonality in this study consistent with previous study findings^24^ and relatively low to moderate level malaria transmission intensity in these regions. Our findings suggest that independent transmission of highly related 622I or diagnostic-resistant *pfhrp2/3-*deleted parasites predominates with bursts of clonal spread. These findings suggest that Ethiopia’s intensive test-and-treat strategies have exerted significant selective pressure on the *P. falciparum* population and are driving rapid expansion of artemisinin and diagnostic-resistant parasites. Though rare, identification of parasites carrying both 622I and *pfhrp2/3* deletion mutations raises concern that parasites with partial resistance to treatment and the ability to escape HRP2-based RDT detection are circulating in Ethiopia.

The presence of *K13* 622I across all sampled districts signals that parasites are under ACT pressure in Ethiopia and indicates that parasites are evolving to escape antimalarial treatment. The 622I mutation was reported previously in two small studies from one site in northern Ethiopia (Gondar), with associated delay in parasite clearance on day 3 of ACT^26^ and increased prevalence over time, from 2.4% in 2014^26^ to 9.5% in 2017-18^27^. While not yet peer reviewed, reports of 622I at high prevalence in Eritrea (16.7% in 2016) and association with 6.3% delayed clearance on day 3 of AL treatment raise further concern about this mutation^33^. The higher prevalence of the 622I mutation in northern Ethiopia (Amhara region) in our study suggests that it originated in northern Ethiopia or Eritrea, though our data are insufficient to determine its origins. The lower frequency of 622I vs. wild-type parasites in polyclonal infections provides evidence that it may decrease fitness within the human host, a consistent trait of artemisinin partial resistance mutations due to loss of function within the *K13* propeller. Taken together, these findings suggest that 622I in Ethiopia represents a meaningful threat to elimination efforts across the Horn of Africa.

As transmission declines in Ethiopia and other settings nearing elimination, the majority of infected individuals are expected to carry single rather than multiple parasite strains. 82% of genotyped samples in our study are monogenomic, consistent with previous findings^24^. The associated increased rate of inbreeding in such settings^34^ is known to favor the spread of drug-resistant strains^35,36^. Decreased parasite competition in low-transmission settings allows strains with resistance mutations that make them relatively less fit in the absence of drug pressure to expand. This is the case for artemisinin partial resistance. We previously showed that false-negative HRP2-based RDT results owing to *pfhrp2/3* deletions are common in Ethiopia and that *pfhrp2* deletion is under recent positive selection^18^. Using a larger MIP panel targeting SNPs across the genome for IBD analysis, we now found that these parasites are closely related and that bursts of clonal transmission appear to be occurring at the district or local scale. These findings support the idea that low transmission and associated parasite inbreeding are important for the expansion of *pfhrp2/3-*deleted populations. This also is consistent with the idea that outcrossing may disrupt co-transmission of *pfhrp2* and *pfhrp3* deletions given they are on separate chromosomes. The rare presence of parasites with both *K13* 622I and *pfhrp2/*3 deletion mutations is not a reassuring finding. Their co-existence in a small number of parasites may simply be a consequence of their distinct origins and insufficient time for the expansion of 622I, *pfhrp2/3*-deleted parasite strains. While combined fitness costs may also play a role in the low prevalence of parasites with both mutations^16,37^, in the absence of inter-strain competition in low transmission settings, there are likely few barriers to the spread of 622I, *pfhrp2/3-* deleted parasites. Our expanded genetic analysis of drug-resistance mutations and parasite population structure confirms that close monitoring of emerging drug- and diagnostic-resistant strains is urgently needed to inform control strategies in the Horn of Africa and neighboring countries.

Different studies suggested that high efficacy of partner drugs (i.e. lumefantrine) impede the spread of ACT resistance in Africa^16,37^. However, we observe high prevalence of mutations associated with resistance to other antimalarial drugs our study, with nearly all genotyped samples carrying the ACT partner drug lumefantrine resistance haplotype (*pfmdr1* NFD)^38,39^ and more than 80% carrying the *pfcrt* N326S background mutation that augments artemisinin partial resistance. No parasites sampled in this study carried other common background mutations observed in SEA (*pffd*-D193Y, *pfcrt*-I356T, *pfarps*-V127M and *pfmdr2*-T484I)^31^. Together, these findings support the need for close monitoring of lumefantrine efficacy and other partner drugs across Ethiopia.

IBD sharing was higher within the *K13* 622I mutant parasite population compared to wild-type parasites, suggesting that 622I mutation emerged or entered into northern Ethiopia in the recent past^27^ and spread to other parts of the country. Highly related parasites are also closely clustered at the district level, a finding expected after clonal transmission. Moreover, our finding of parasites with high IBD and low overall COI in this study indicates low ongoing transmission across the three regions and that most recombination is between highly-related or clonal strains^40,41^. IBD analysis also showed high relatedness and clonal expansion of *pfhrp2/3* double-deleted parasites (most likely not detected by HRP2*-*based RDTs) at the local scale, with distinct populations of very closely related *pfhrp2/3-*deleted parasites observed in several districts. Clonal spread with local inbreeding could facilitate rapid spread of *pfhrp2/3-*deleted parasites that are expected to escape diagnosis by RDTs. Our data also reveals higher prevalence of 622I mutation among *pfhrp2/3* non-deleted compared to double-deleted parasites, a finding that might be seen when *pfhrp2/3* deletion leads to misdiagnosis, leaves patients untreated, and results in *pfhrp2/3-*deleted parasites exposed to less ACT pressure. Supporting this idea, we observed more frequent co-occurrence of *pfcrt-* K76T mutation suggestive of empirical chloroquine treatment for presumed non-falciparum malaria.

Our study is not without limitations. First, travel histories from malaria cases and samples from neighboring countries are not included and thus tracking resistant strain importation is not addressed in detail. Second, the parent study was designed to evaluate RDT failure and could introduce selection bias, including under sampling of low-parasitemia and submicroscopic infections or oversampling of monogenomic infections. We therefore adjusted our *K13* 622I prevalence estimates to improve the generalizability of our findings. Third, the areas studied represent regions with relatively higher transmission (Amhara, Gambella and Tigray) and do not include other parts of the country, making it difficult to extrapolate our findings across the country. It may be that other regions have lower prevalences of drug and diagnostic resistance mutations, or that prevalences are even higher in lower transmission settings. Further study within Ethiopia and surrounding countries is warranted.

Overall, our study provides evidence that the ongoing selective pressures exerted on parasite populations in Ethiopia by HRP2-based RDT diagnosis^42^ and ACT treatment^43^ could facilitate co-occurrence of diagnostic and drug resistance, representing a double threat to malaria elimination. Ethiopia’s recent transition to alternative RDTs may reduce selective pressures favoring *pfhrp2/3-*deleted strains. However, we observe 622I mutation in multiple regions alongside *pfhrp2/3*-deleted parasites, and concerning examples of co-occurrence that could yield parasites resistant to both diagnosis and treatment. As Ethiopia and other countries in the Horn of Africa approach malaria elimination, diagnostic and drug resistance may be more likely to co-occur. These findings also illustrate the value of targeted parasite genomic analysis as part of large-scale malaria surveys and demonstrate the need for close monitoring of ACT efficacy that includes advanced molecular surveillance in Ethiopia.

## Materials and Methods

### Study sites and sample genotyping

A total of 920 samples from three regions (Amhara = 598, Gambella = 83 and Tigray = 239) (**Supplementary Figure S1**) previously assessed for *pfhrp2/3* deletions^18^ were further genotyped using molecular inversion probes (MIPs). Sampling strategy, samples collection, DBS samples transportation, DNA extraction and initial molecular analysis were described in detail in our previous study^18^. The parent study was approved by the Ethiopian Public Health Institute (Addis Ababa, Ethiopia; protocol EPHI-IRB-033-2017) and the World Health Organization Research Ethics Review Committee (Geneva, Switzerland; protocol ERC.0003174 001). Parasite sequencing and analysis of de-identified samples was deemed nonhuman subjects research by the University of North Carolina at Chapel Hill (NC, USA; study 17-0155).

### MIP capture, sequencing and variant calling

DNA originally isolated from DBS samples were captured and sequenced using two separate MIP panels: (1) a drug resistance panel (n = 814) designed to target mutations and genes associated with antimalarial resistance, and (2) a genome-wide panel (n = 1832) designed to target SNPs to evaluate parasite connectivity and relatedness (**Supplementary Data 1** and **2**)^28,44^. MIP capture and library preparation were performed as previously described^17^. Sequencing was conducted using an Illumina NextSeq 550 instrument (150 bp paired-end reads) at Brown University (RI, USA).

The MIPtools (v0.19.12.13; https://github.com/bailey-lab/MIPTools) bioinformatic pipeline was used for processing of sequencing data and variant calling. Briefly, this pipeline employs MIPWrangler software to stitch paired reads, remove sequence errors, and predict MIP microhaplotypes leveraging the unique molecular identifiers (UMIs) in each arm. The haplotypes for each target were mapped to the *P. falciparum* 3D7 reference genome (PlasmoDB-42_Pfalciparum3D7) using Burrows-Wheeler Aligner (BWA)^45^ and variant calling was performed on these samples using freebayes^46^. Downstream analyses were performed on generated variant calling files (VCF) as well as translated tables based on 3D7 transcriptome for coding mutations. For genome-wide MIP panel, variants were quality filtered by removing those with less than three UMIs within a sample and less than 10 UMIs across the entire population. The drug resistance panel included known SNPs in *pfcrt, pfdhfr, pfdhps, pfmdr1, K13* and other putative drug resistance genes and has been described elsewhere^28^ (**Supplementary Data 2**). First prevalence was calculated as (p = m/n*100, where p = prevalence, m = number of infections with mutant alleles, n = number of successfully genotyped infections) (Table S3). Unweighted prevalence was calculated using the miplicorn R package version 0.2.90 (https://github.com/bailey-lab/miplicorn) and vcfR R package version 1.13.0^47^. Mutant combinations were plotted and visualized using *UpSet* Package in R version 1.4.0^48^. Because dried blood spot sampling differed based on RDT results (participants with HRP2-/PfLDH+ results were purposefully oversampled for molecular characterization in the parent study), we adjusted our *K13* 622I prevalence estimates by weighting for the relative sampling proportions of RDT-concordant (HRP2+) and discordant (HRP2-/PfLDH+) samples. This was achieved by weighting RDT profile-specific prevalence estimates by the total number of *P. falciparum-*positive individuals presenting with that RDT profile in the parent study by district, region, and overall. Finally, 95% confidence intervals for these weighted prevalence estimates were estimated using bias corrected and accelerated (BCa) bootstrapping (n = 2000 replications for district and region-level estimates, n = 3000 replications for overall study estimate) using the R packages boot (version 1.3-28) and confintr (version 0.2.0). Mutant combinations were plotted and visualized using *UpSet* Package in R version 1.4.0^48^. For the genome-wide MIP panel, only biallelic variant SNPs were retained for analysis. Genome positions with more than 50% missing data (**Supplementary Figure S3A)**, and samples missing 50% of sites (**Supplementary Figure S3B)** were removed leaving 609 samples and 1395 SNPs from the genome-wide panel **(Supplementary Figure S4A)**, which are distributed across 14 *P. falciparum* chromosomes (**Supplementary Figure S4B**). The drug resistance panel includes SNPs across known *P. falciparum* drug resistance genes that have been described elsewhere^28^.

### Complexity of infection analysis (COI)

To estimate the COI, we used THE REAL McCOIL R package categorical method^49^. As DBS sampling in the parent study favored RDT discordant samples (HRP2-/pfLDH+) and could bias our COI estimates, we estimated overall and district level prevalence of monogenomic infections by weighting for the relative sample proportions of RDT concordant and discordant samples in the parent survey. We also calculated within-host fixation index (F_ws_) using R package moimix 2.9^50^ as another measure of within-host diversity of the parasites, which measures the probability that any random pair of infections carry different alleles at a specific locus. It was calculated for each infection as follows, F_ws_ = 1-(Hw/Hs), where Hw is the infection heterozygosity across all loci and Hs is the heterozygosity of the population from which the infection was sampled. As F_ws_ calculation based on the frequency of alleles per individual compared to that within the source population, it allows comparison between populations. F_ws_ range from 0 to 1, the sample was classified as having multiple infections (polyclonal) if F_ws_ < 0.95 and monoclonal (single strain) infections if F_ws_ ≥ 0.95. Samples with F_ws_ < 0.95 were considered to come from mixed strain infections, indicating within-host diversity.

### Population structure and genetic differentiation

To assess whether parasite populations within Ethiopia clustered based on their geographic origin, or *pfhrp2/3* deletion status, we first conducted principal component analysis (PCA) using SNPRelate R package version 1.30.1^51^. The eigenvalues generated from filtered VCF using snpgdsPCA function used as input file and the result was visualized using ggplot2 R package version 3.4.0. We calculated pairwise genetic differentiation (*F*_ST_) as a measure of genetic divergence between populations using PopGenome R package version 2.7.5^52^.

### Analysis of parasite relatedness using Identity-by-Descent (IBD)

To measure relatedness between *P. falciparum* parasites and identify regions of the genome shared with recent common ancestry, the inbreeding_mle function of the MIPAnalyzer software version 1.0.0 was used on monogenomic samples to calculate IBD^44^. We determined IBD sharing variation at regional and local scale (district level) to assess spatial patterns of parasite connectivity and transmission dynamics at micro-local level comparing deleted and mutant parasites vs wild-type. Networks of highly-related parasites per *K13* 622I mutation status or *pfhrp2/3* deletion status were generated using the igraph R package version 1.3.5^53^.

## Data Availability

All data produced in the present study are available upon reasonable request to the authors.

## Acknowledgments

We thank the EPHI research teams for conducting the fieldwork during the parent study. We would also like to thank all of the participants and family members who contributed to this study.

## Data Availability

All sequencing data available under Accession no. pending at the Sequence Read Archive (SRA) (pending), and the associated BioProject alias is pending.

## Author contributions

AAF, JBP, JJJ and JAB conceived the study. SF led the parent study, with contributions from MH, BG, HM, BP, SH, JC, and JBP. AAF, CR, CH performed laboratory work. AAF led genetic data analysis and wrote the first draft of the manuscript. JBP, JJJ and JAB supported genetic data analysis and interpretations of results. All authors contributed to the writing of the manuscript and approved the final version before submission.

## SUPPLEMENTARY MATERIAL

### Supplementary Tables

#### Supplementary tables are compiled into a single file for ease of viewing

**Table S1**. Prevalence of monogenomic infections per district weighted by RDT discordant prevalence.

**Table S2**. Weighted prevalence of key drug resistance mutations per district. Weighting calculated by RDT profile (see Methods).

**Table S3**. District level prevalence (unweighted) of all nonsynonymous mutations across different drug resistance genes. The prevalence of antimalarial resistance mutations detected by the MIPs is shown for each geographic location. Mutations above 1% prevalence in any of the districts shown. AA = mino Acid; n = number of samples genotyped; m = number of samples carry mutant allele.

### Supplementary Data

#### Supplementary tables are compiled into a single file for ease of viewing

**Data 1**. List of loci included in genome wide MIP panel.

**Data 2**. List of loci included in drug resistance MIP panel.

**Data 3**. Metadata for 609 successfully sequenced samples using the genome wide MIP panel. This data was used for COI estimation and population genetic analysis.

**Data 4**. Metadata and genotype file for all successfully sequenced samples (varies per marker) using the drug resistance MIP panel. Only successfully sequenced samples were used for drug resistance marker prevalence estimates. 0 = Reference Call, 1 = Alternative heterozygous call, 2= Alternative homozygous call, -1 = Missing call.

## Supplementary Figures

**Figure S1.**
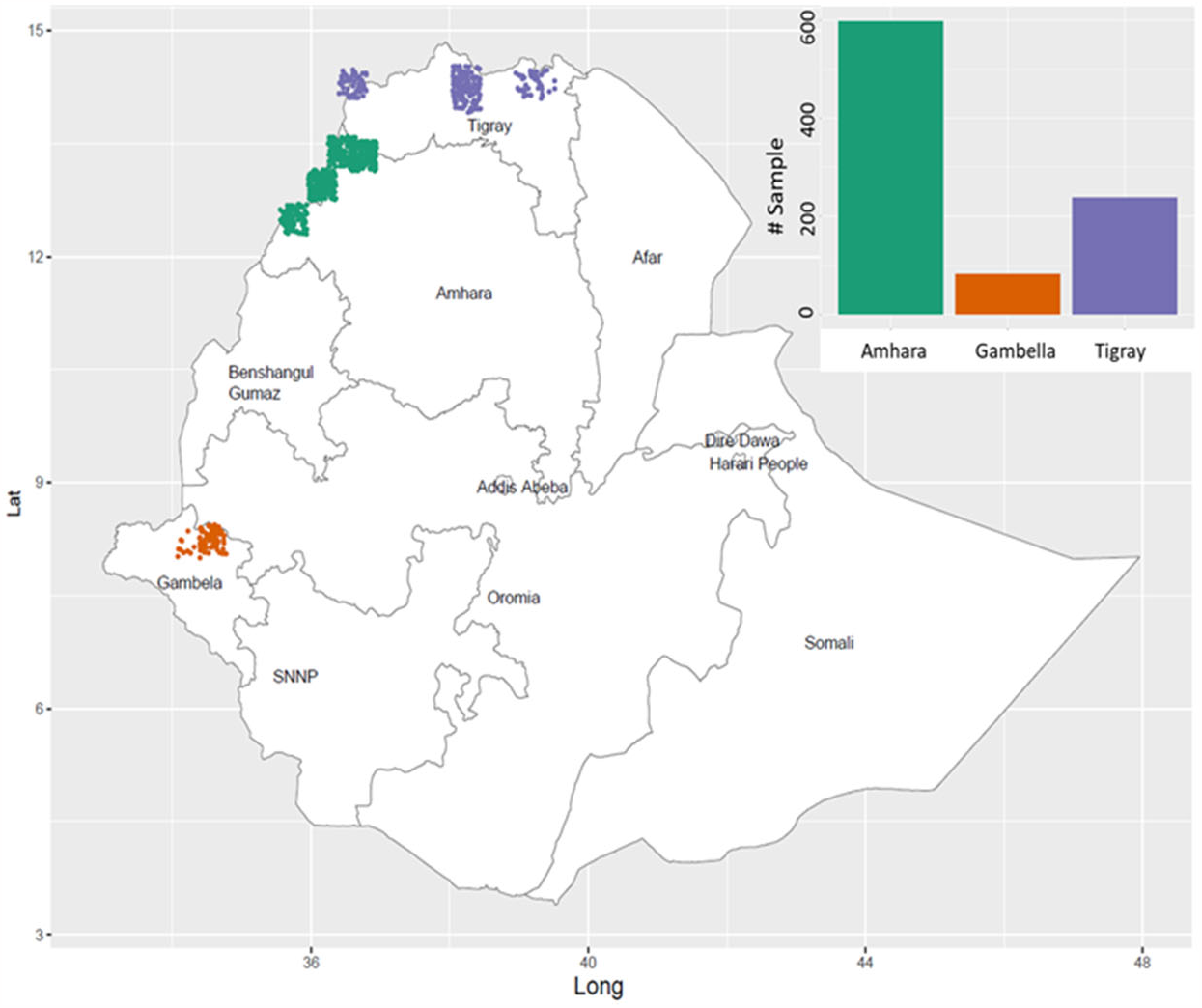
Description of sequenced samples. Spatial distribution of sequenced samples at district level (color dots in map) and regional level (color bar plot). Colors indicate regions.

**Figure S2.**
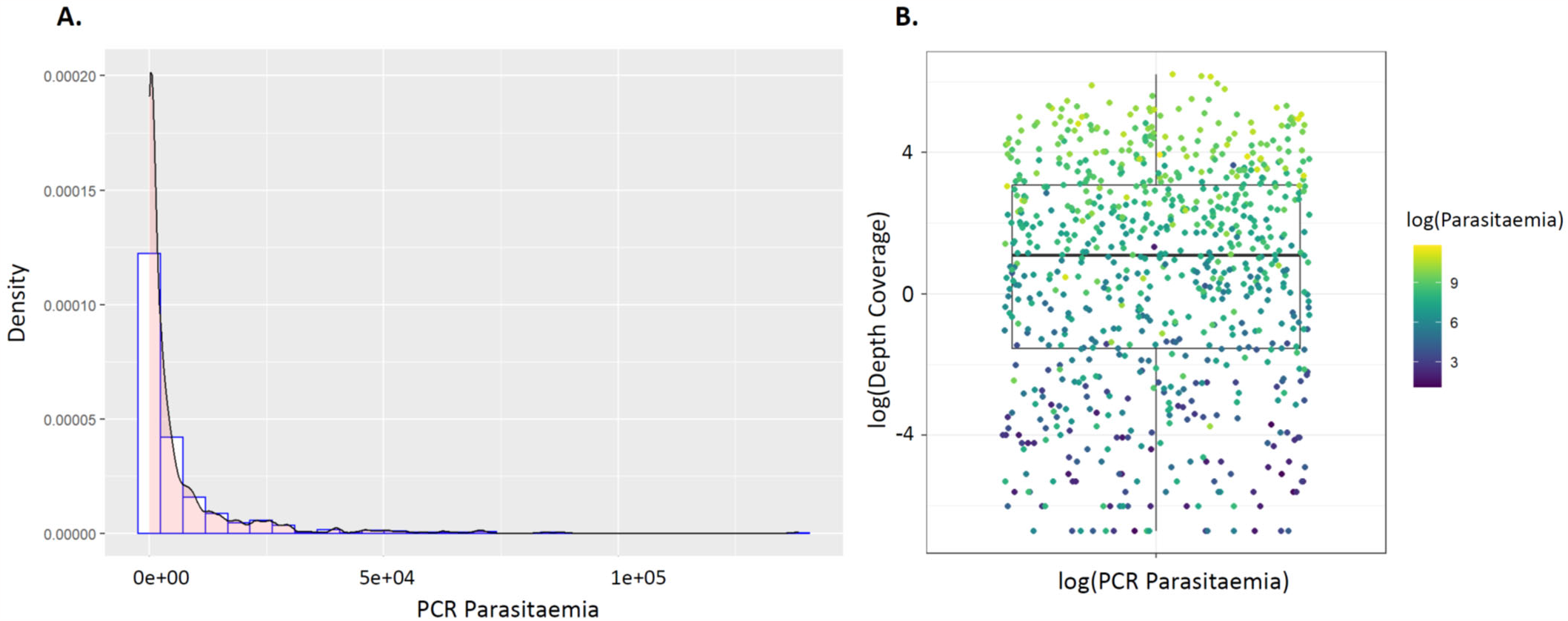
PCR parasitemia distribution and association between sequencing coverage and parasitaemia. **A)** Density plot showing parasitemia distribution with median parasitemia = 1411 parasite/ul. **B)** Association between sequencing coverage and parasitaemia. The MIP sequencing success is parasitaemia dependent as shown in the heatmap color.

**Figure S3.**
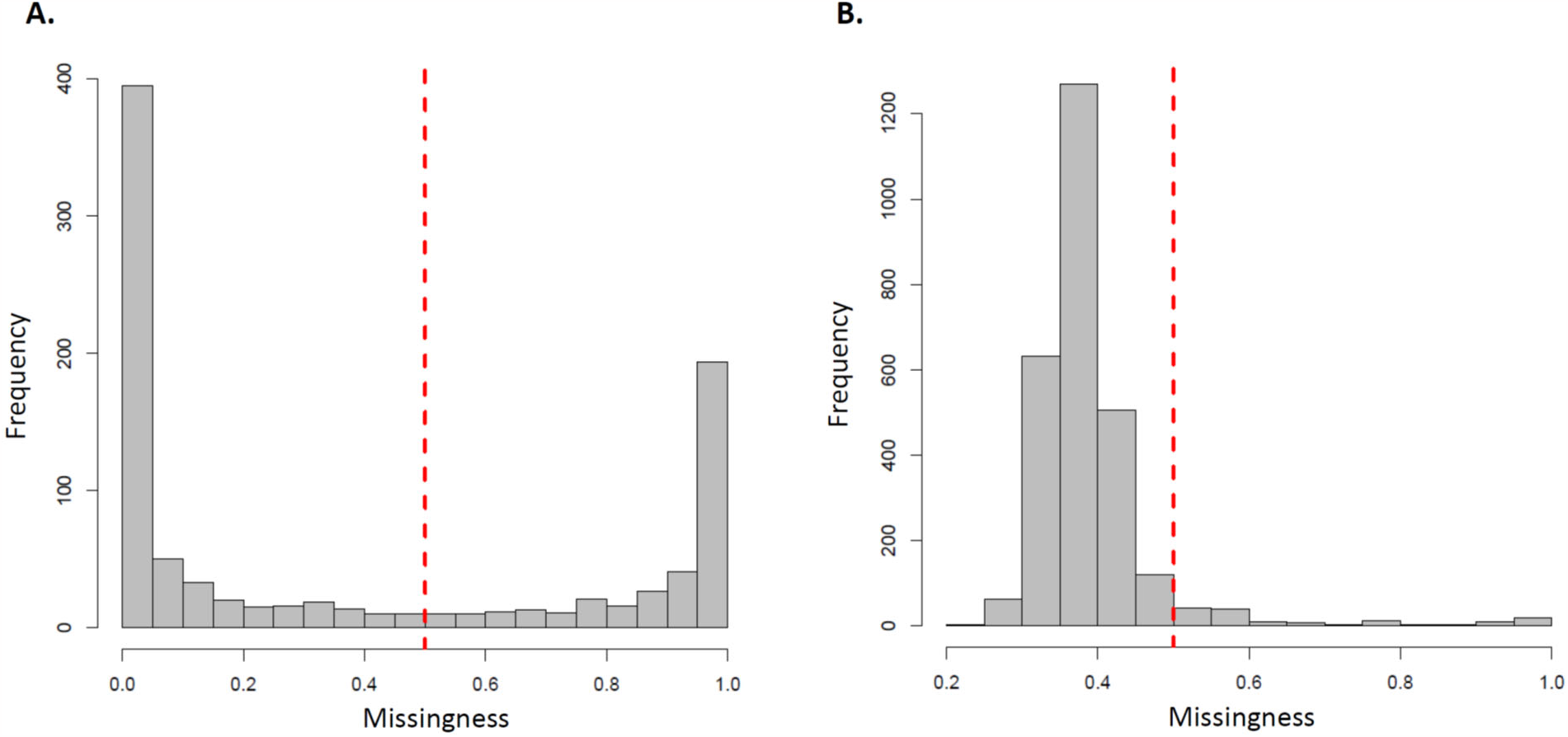
Sample (A) and SNP (B) missingness. **A)** Samples with >50% low-coverage loci were dropped as shown broken read line. **B)** Variant sites were then assessed by the same means in terms of the proportion of low-coverage samples, and sites with >50% low-coverage samples were dropped. Broken read line shows 50% threshold criteria we used to remove samples and loci from downstream analyses.

**Figure S4.**
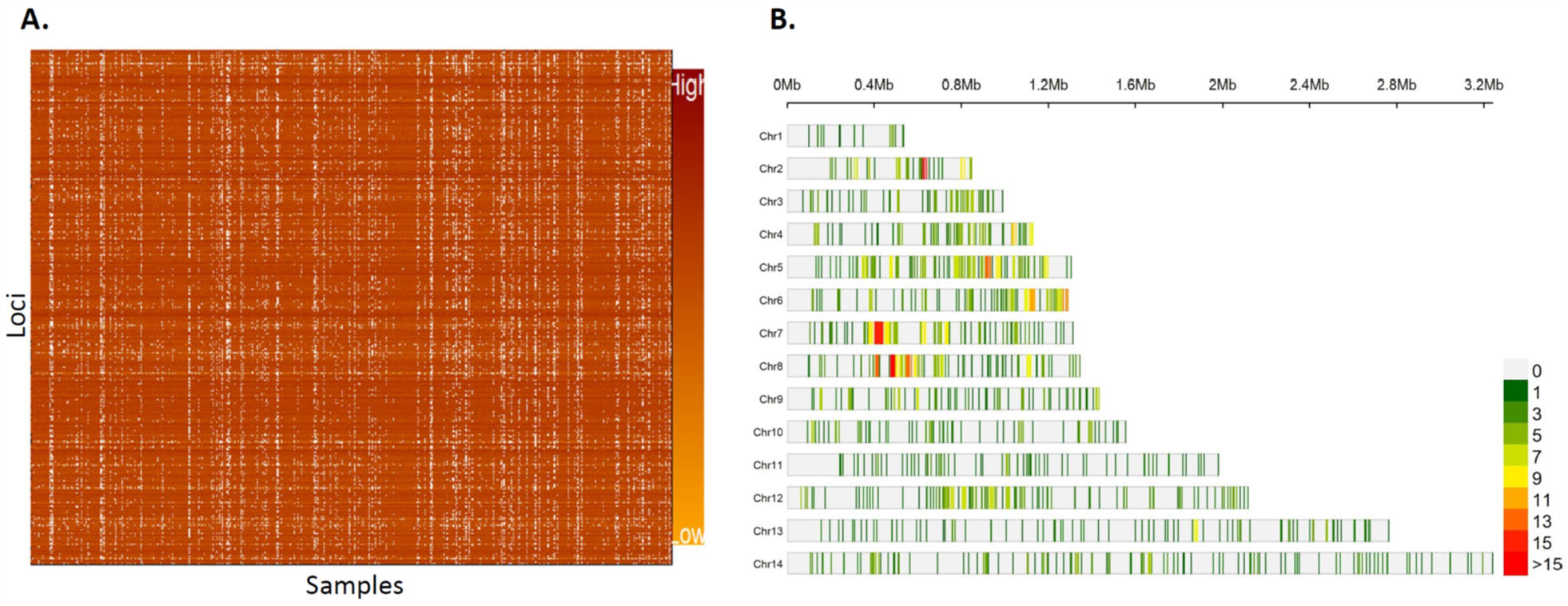
Successfully sequenced samples across three regions in Ethiopia and retained genome wide loci. **A)** Association between sequencing coverage and parasitaemia. The MIP sequencing success is parasitaemia dependent as shown in the heatmap color. **B)** Distribution of retained SNPs across *Plasmodium falciparum* chromosomes. The plot shows distribution of 1395 retained high quality biallelic SNPs across the 14 *P. falciparum* chromosomes within 0.025 Mb window size. Color coded from light gray for masked regions with no SNPs to red for regions containing high number SNPs per chromosome.

**Figure S5.**
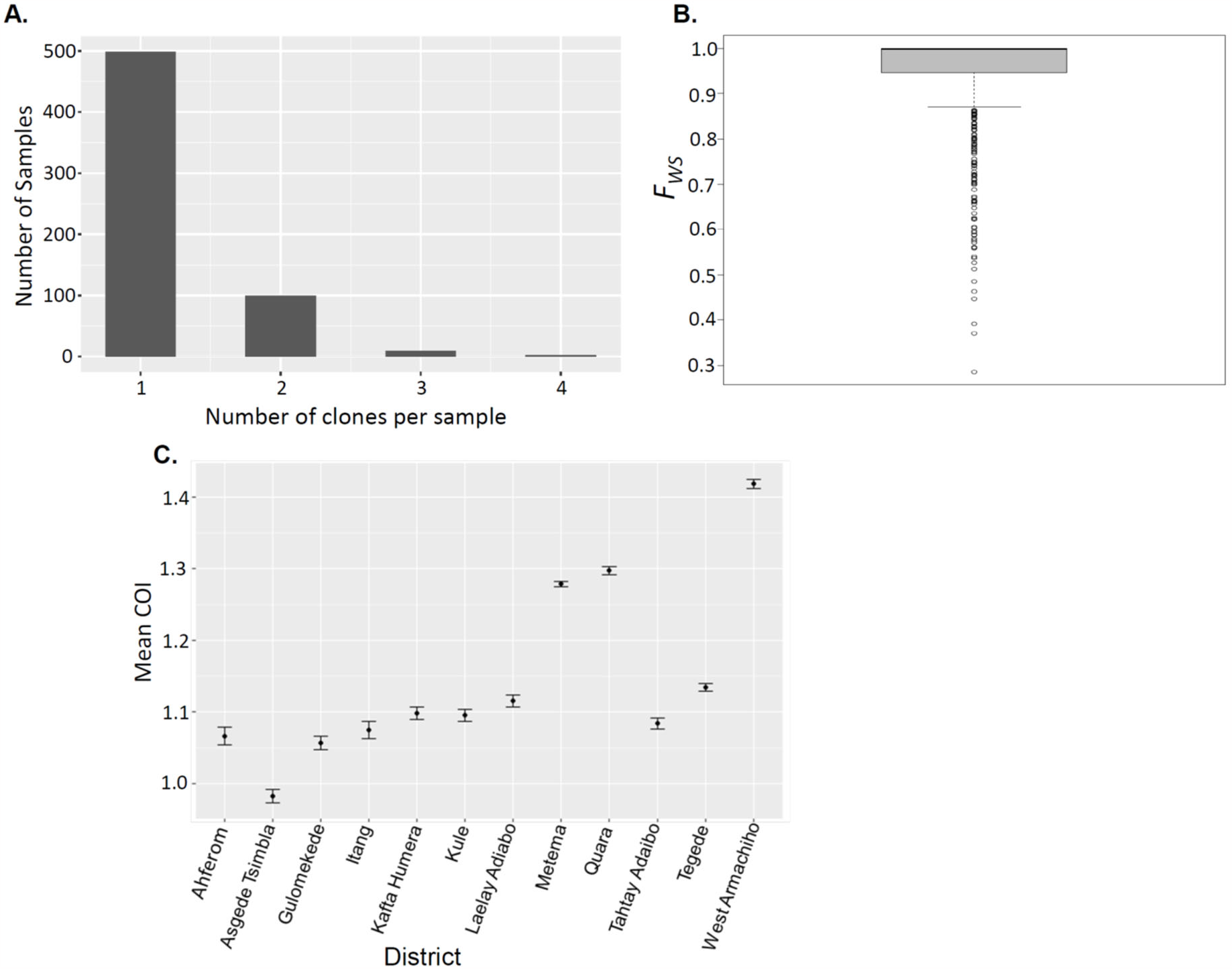
Complexity of infections. **A)** Distribution Number of clones per sample across genotyped samples showing most of isolates carrying one clone (COI=1). **B)** Cumulative within-infection FWS fixation showing majority of isolates classified as monogenomic (FWS > 0.95). Number of clones per sample. **C)** Spatial heterogeneity of mean complexity of infections per district across three regions in Ethiopia. Vertical lines show 95% confidence intervals.

**Figure S6.**
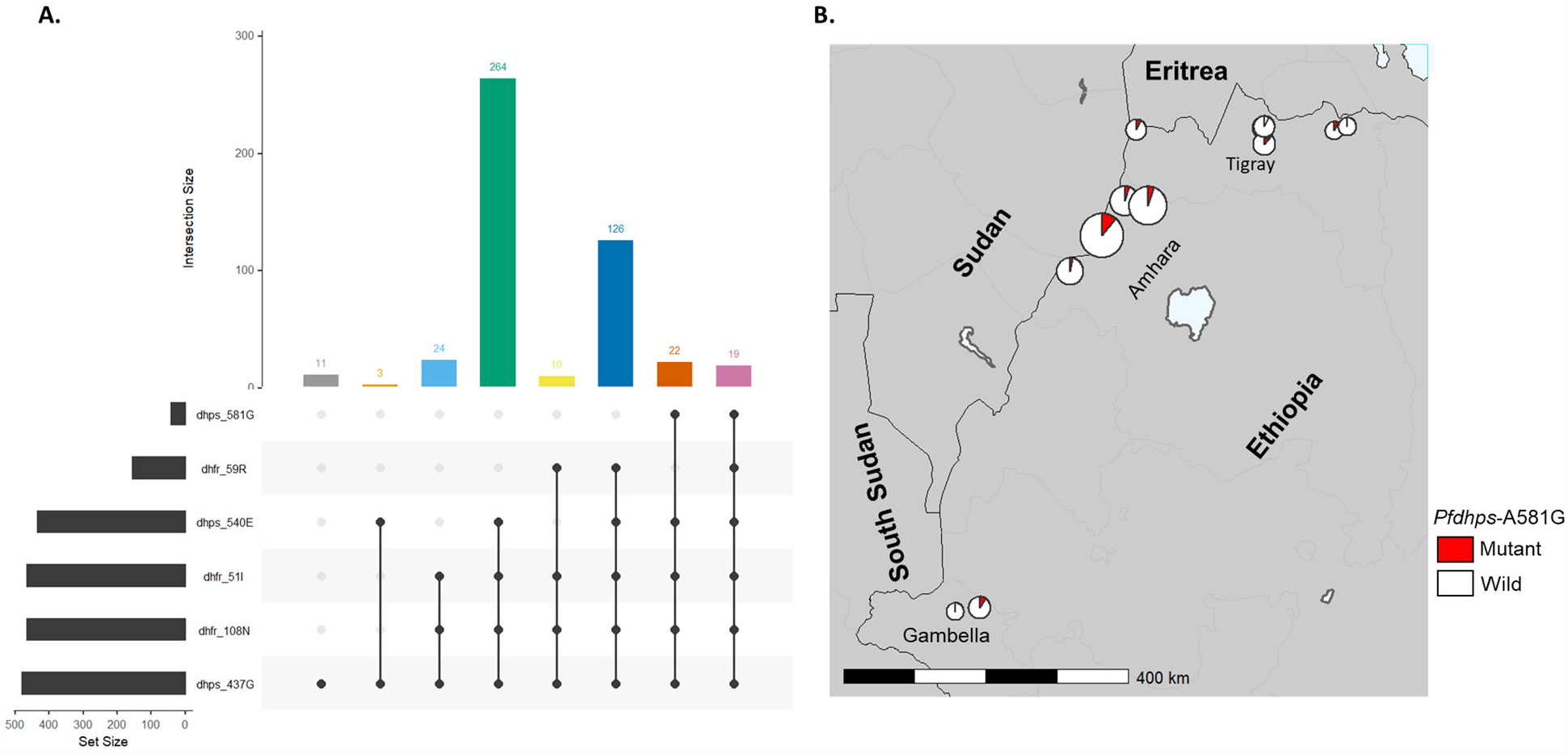
Prevalence of *Pfdhfr* and *Pfdhps* mutations across Ethiopia. **A)** UpSet plots showing the number of times each combination of mutations was seen for *Pfdhfr* and *Pfdhps*. **B)** Spatial distribution of *Pfdhps A581G* mutation at district level. Colors indicate mutation status and size of pie chart is proportional to sample size per district.

**Figure S7.**
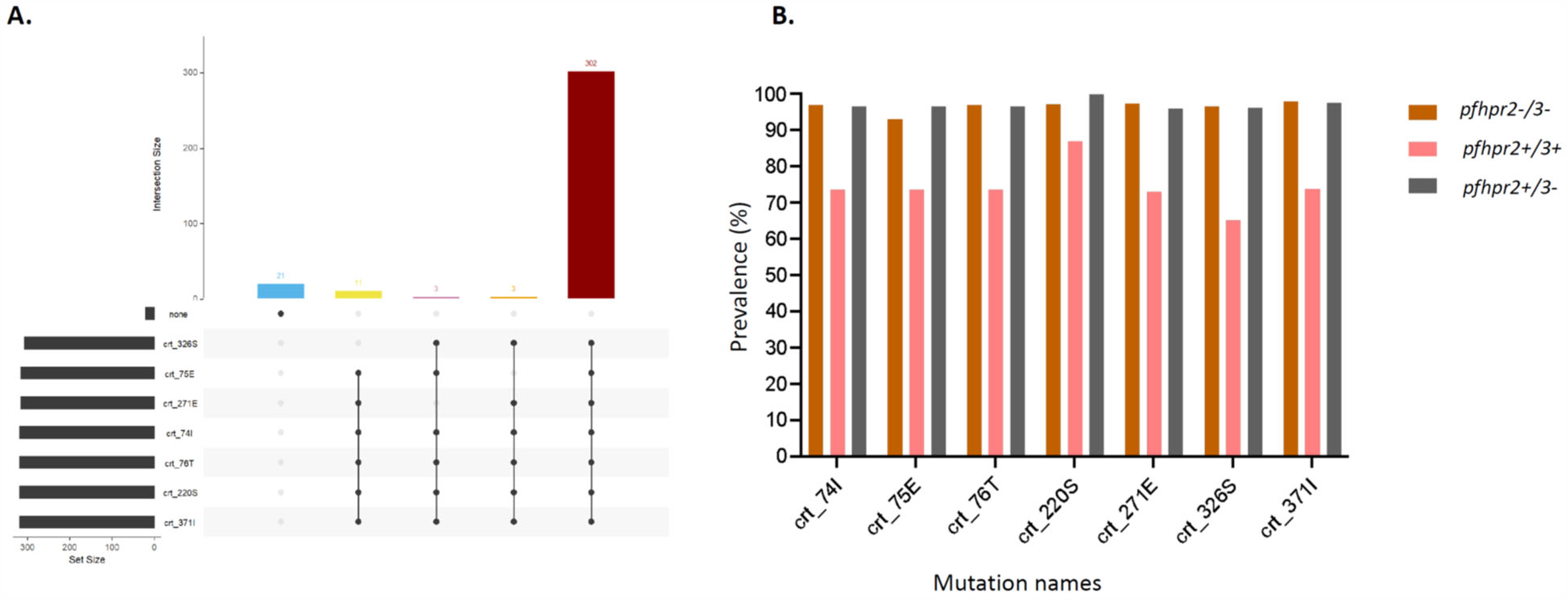
Prevalence of *pfcrt* mutations. The UpSet plot shows the number of times each combination of mutation was observed within ***pfcrt*** (A), and prevalence of these mutations by *pfhrp2/3* status (B).

**Figure S8.**
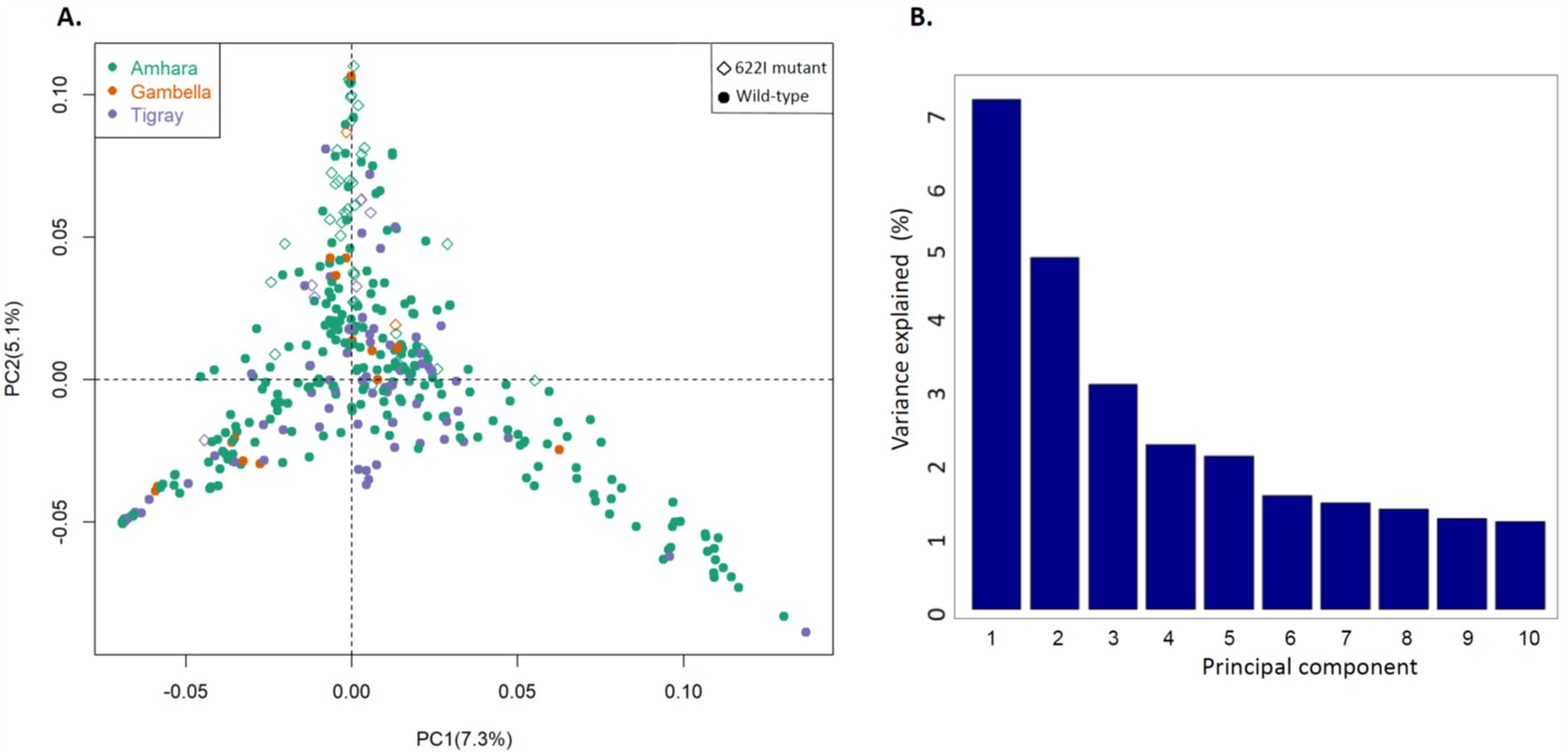
Population structure of *P. falciparum in* Ethiopia. **A)** Principal component analysis *P. falciparum* populations per region. Colors indicate sample origin and shape indicates *K13* 622I mutation status (circle indicates wild and diamond indicates mutant). Percentage of variance explained by each principal component presented (%). **B)** Percent of overall variance explained by the first 10 principal components during PCA.

**Figure S9.**
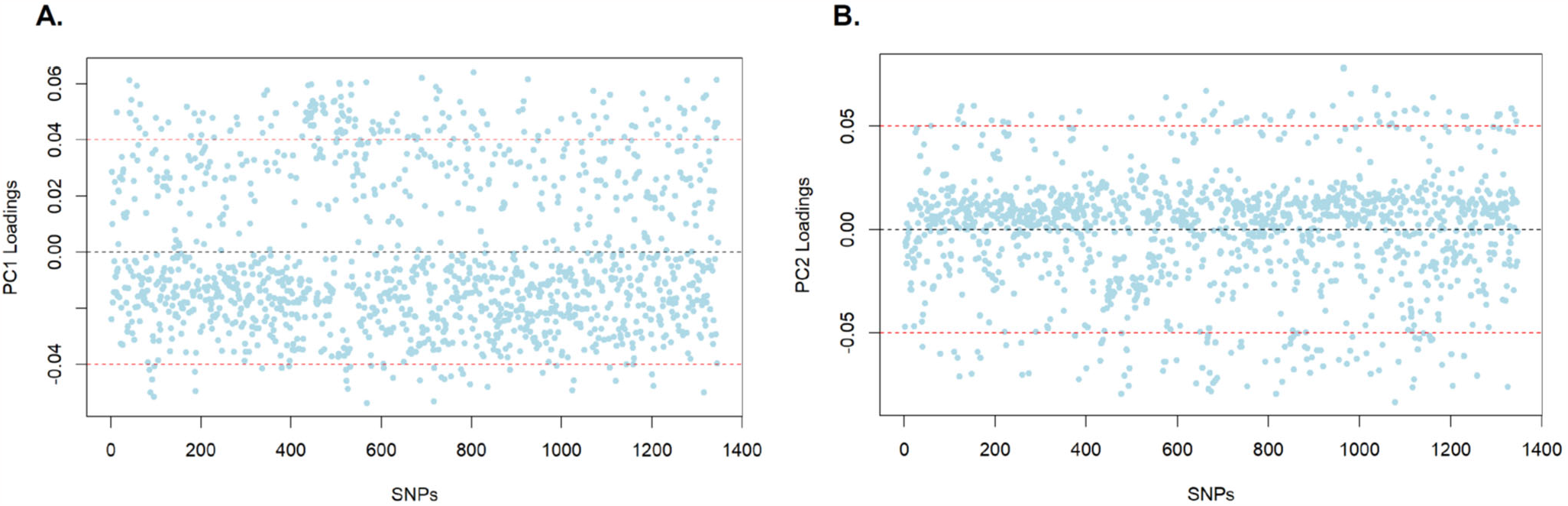
PCA loading values. PC1 (A) and PC2 (B) are shown by SNP. Cutoffs show SNPs that highly contribute to positive or negative distribution of samples in the PC plots.

